# Barriers and facilitators to sexual and reproductive health rights for Persons with Disability in Nepal: a scoping review

**DOI:** 10.1101/2023.04.19.23288803

**Authors:** Sanju Bhattarai, Pratap KC Saugat, Sampurna Kakchapati, Shraddha Poudel, Sushil Chandra Baral, Cicely Marston

## Abstract

Persons with disability have the same sexual and reproductive health needs as people without disability but their rights have consistently been overlooked. They face numerous challenges to access sexual and reproductive health services in Nepal, however coherent evidence on nature, size, and extent of these challenges are not available. We carried out a literature review to explore barriers and facilitators encountered by persons with disability while accessing sexual and reproductive health services in Nepal. We reviewed published government policies on reproductive health and disability, searched PubMed database and used google scholar search engine to find literature published between 2011 to 2021 that reported on barriers and facilitators to sexual and reproductive health rights for person with disability in Nepal. Out of 2145 identified literature only 21 literatures meeting the eligibility criteria were included in the analysis. We found inadequate inclusion of PWDs in health sector policies, lack of knowledge about SRH needs, misconception and poor attitude and lack of social support in accessing SRH rights and services. In Nepal, people with disabilities face multitude of barriers in accessing sexual and reproductive healthcare. Multilevel measures informed by further studies on vulnerabilities and experience of different subgroups of PWDs.

## Introduction

Worldwide 15% of the adult population (15 years and older) lives with some form of disability, the proportion is higher (18%) in low and middle income countries(1) and estimates from Nepal range between 4.0%(2) to 13.6%(3). Disability is an important social determinant of health(4): person with disabilities (PWDs) experience lower educational attainment(5), less income(6), and worse health outcomes(7) compared with people without disabilities. These circumstances reduce access to health and social services forcing many PWDs into poverty, social exclusion and marginalization(1).

### Sexual and Reproductive Health needs of PWDs

Persons with Disabilities have the same SRH needs as people without disability but their rights have consistently been overlooked and ignored(8). They are often deprived the rights to establish relationships, marry or to start a family, in extreme cases they are even forced into sterilization, abortion and unwanted marriages(9). PWDs are restricted from accessing health care for reasons including physical inaccessibility of facilities and infrastructure, unavailability of suitable and affordable transportation, inappropriate communication materials, lack of assistive devices, and long waiting times(10). Other factors that drive poor access to SRH services are health care providers’ negative attitudes and ignorance about the health needs of the PWDs, and lack of insurance or safety net to cover healthcare costs(8). These barriers can deprive PWDs of basic SRH information. Lack of knowledge and skills in partner negotiation increases the likelihood of engaging in high-risk behaviors(11). Even when able, PWDs access and use SRH services less frequently(12,13) increasing the risk of being infected with Human Immunodeficiency virus (HIV) and other sexually transmitted infections (STIs)(14). Incorrect assumptions and beliefs about the sexual drive of PWDs, considering them either sexually inactive or hyperactive, can also reduce their access to SRH services increasing their exposure to abuse and adverse health consequences(15,16). PWDs are at increased risk of experiencing abuse and gender-based violence, compared to others, they are three times more likely to become victims of physical and sexual violence, higher among those financially and physically dependent on the perpetrator(8). A meta-analysis reported that sexual victimization is higher among women with disabilities.(17)

These vulnerabilities underscore that the PWDs require SRH education and care as much as, or possibly even more than, people without disabilities. However, their SRH rights have been largely ignored by both the disability community and those working on SRH(8). Universal access to SRH is a fundamental human right as emphasized in the Sustainable Development Goals on good health and wellbeing and gender equality(18). Within this context, an understanding of the factors affecting access to SRH information and services by PWDs is urgently needed. This review documents the barriers and facilitators for tailoring SRH programmes to meet the specific needs of the PWDs in Nepal.

### Policy environment for PWDs in Nepal

As a signatory to several international human rights instruments including the Universal Declaration of Human Rights’ 1980 and the United Nations Convention on the Rights of PWDs (CRPD) 2006, the Nepal government is obliged to ensure the rights of PWDs. Over the years Nepal has developed several acts, namely, Protection and Welfare of the Disabled Persons Act, 1982 and the, the Education Act 1992, Child Rights Act 1992 and Local Self-governance Act 1999. However, ‘The 2013 constitution of Nepal’ set out specific commitments to ensure the basic human rights of its citizens, specifically mentioning to uphold the rights of PWDs, and moved the needs of PWD higher up the national development agenda. In 2017 Nepal developed an act on ‘Rights of the PWDs upholding the eight principles of the United Nations Convention on the Rights of Persons with Disabilities (CRDP) (respect and dignity, non-discrimination, full and active participation, respect and accepting PWDs, equal opportunity, accessibility, equality between gender, respect for children with disability)’. These policy frameworks set out government obligation to effectively manage disability, and the consecutive national plans increasingly provide for health and community-based services for PWDs such as rehabilitative schemes, special needs education, and grants and social security. The Ministry of Women, Children and Senior Citizen is largely responsible for implementing these programs. Within the health sector, the National Health Policy 2014 mandated universal and equitable health services, including health care for PWDs under the basic health services, allocating some funds for treatment of PWDs patients(19). Along with general health services, article 25 of the CRPD also urged member states to ensure the sexual and reproductive health (SRH) rights of PWDs, through provision of quality health care including SRH as provided to other people(20). Nepal’s Constitution 2015(21) vows to guarantee the SRH rights of its citizens including PWDs. Despite protection of SRH being a constitutional rights, the PWDs continue to face social injustice and are denied their right to equitable health services(22). Estimates show more than 40% of PWDs in Nepal do not have access to necessary healthcare service(23). The paucity of published articles on SRH of PWD limited our ability to make a coherent argument on the barriers and facilitators faced by different subgroups of PWDs in Nepal. Therefore, we aimed to identify and synthesize the current literature on perceived barriers and facilitators to SRH among PWDs in Nepal and the interventions to address their needs.

### Theoretical Framework

We used the Kaufman et al.’s model on developing a socio-ecological framework(24) to guide organization and presentation of the results. The model provides a combination of multilevel factors individual (e.g. beliefs, behaviors), social and community (e.g. norms), institutional and health system (e.g. health services, education), and structural (e.g. laws, safety, social protection) factors that may interact in ensuring SRH rights of PWDs.

## Methods

We carried out a literature review to answer three research questions:

1. What are the barriers and facilitators to SRH for PWDs in Nepal?
2. What evidence is there relating to SRH interventions for PWDs in Nepal?
3. Are PWDs included in SRH policies and guidelines in Nepal?

### Search Strategy

For the purpose of this review the definition of PWDs used was ‘‘long term physical, mental, intellectual, or sensory impairments which, in interaction with various barriers, may hinder their full and effective participation in society on an equal basis with others’’(20) and for SRH the International Conference on Population and Development (ICPD) definition that includes maternal health, newborn health, family planning, STIs, including HIV, and gender-based violence(25) was used. These definitions guided finalization of key search terms.

On 9th of March 2022, we searched the PubMed database to retrieve articles. Our search terms included MESH terms (presented in the supplementary table 1) including search domains “Sexual and Reproductive Health’, ‘Disability’, and ‘Nepal’. In addition, we used publish or perish software(26) to search for articles using google scholar search engine using search terms “Sexual and Reproductive health”, “People with Disability” and “Nepal”. To understand the extent to which PWDs are included in SRH policies (research question 3), we hand searched websites of organizations working for PWDs, government reproductive health and disability related policy documents, guidelines, national surveys, and routine monitoring data from Nepal.

### Eligibility criteria

We included original articles, case studies, systematic reviews, and grey literature that published qualitative or quantitative findings relevant to PWD and SRH from Nepal between August 2011 to August 2021, this was done to gain an understanding of the research landscape with a greater precision during this period. We included all publications reporting on any of the SRH outcomes in the ICPD definition mentioned earlier.

### Screening

Two researchers (SPK and SP) screened the title and abstract of the articles. Those articles not mentioning barriers or facilitators or SHR outcomes and interventions for PWDs were excluded.

### Data extraction and analysis

We extracted information from the included articles on study characteristics (e.g., publication year, study population, study aim, design), and barriers, facilitators, and intervention characteristics. Similarly, we extracted information on ownership, inclusion of PWDs and specific activities for PWDs from policy documents. Two researchers (SPK and SP) extracted the information, and any disagreements were resolved by consensus after discussing with third researcher (SBW).

## Results

Fig 1 shows the screening process of the published articles. Altogether, 2145 articles were identified across PubMed (1147) and Google scholar (998). We removed 492 duplicates. Next, we screened the title and abstract of the 1653 remaining articles, 1631 were excluded for not reporting on any of the SRH outcomes from Nepal. The remaining 21 articles were included in this review.

**Fig 1.**
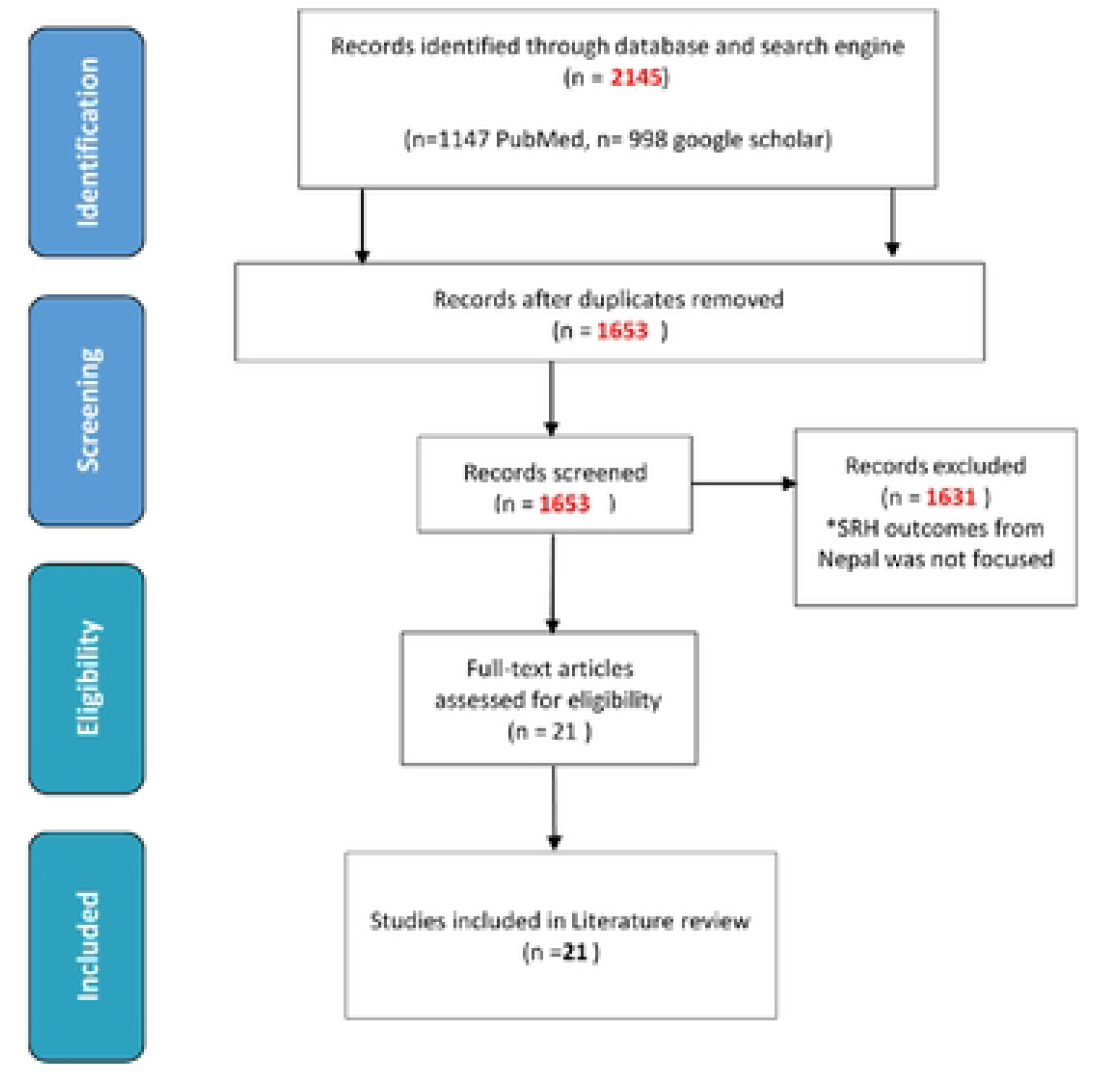
PRISMA flow chart of published articles reviewed

### Characteristics of the publications included

Table 1 shows the characteristics of the 21 articles included in the review. Sixteen were peer reviewed journal articles (6 mixed method, 4 qualitative and 3 quantitative, 1 systematic review, 1 Intervention and 1 Randomized Control Trial) and the remaining five were: 2 policy papers, 1 situational analysis report and two case studies. Out of the 16 peer reviewed articles, 14 reported on female PWDs, two on both male and female PWDs, one each reported care takers and health care provider’s perspectives. Thirteen of the studies were conducted in rural setting and three in both urban and rural settings and for the rest not mentioned. One of the studies recruited participants from health facility and another one from refugee camp. Sixteen of the 21 studies explored facilitators and barriers to SRH issues.

**Table 1:**
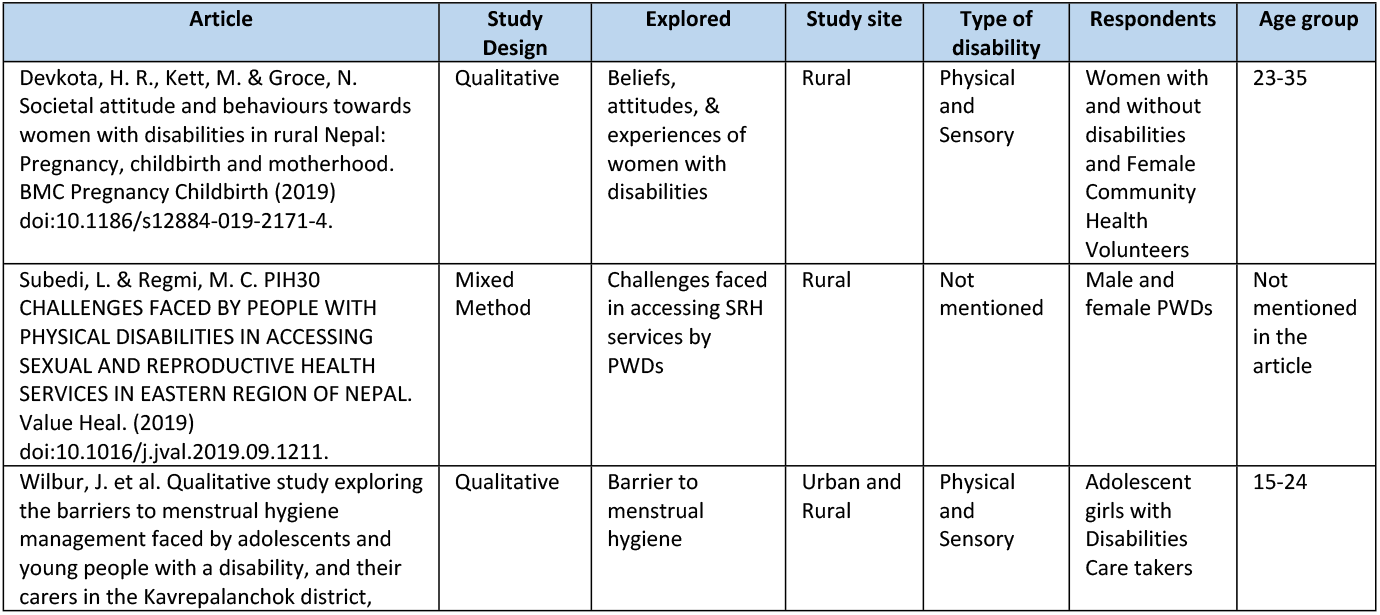
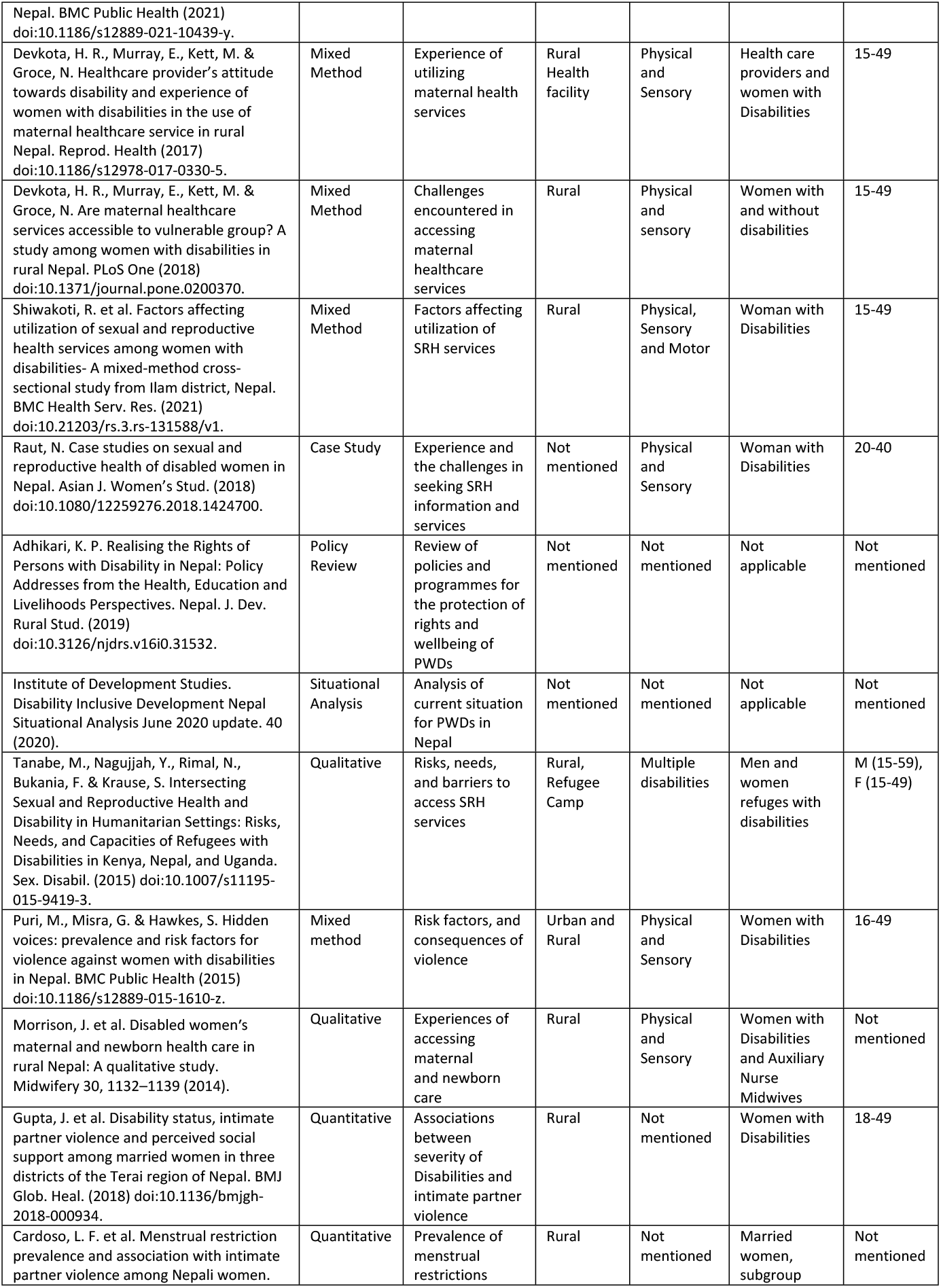
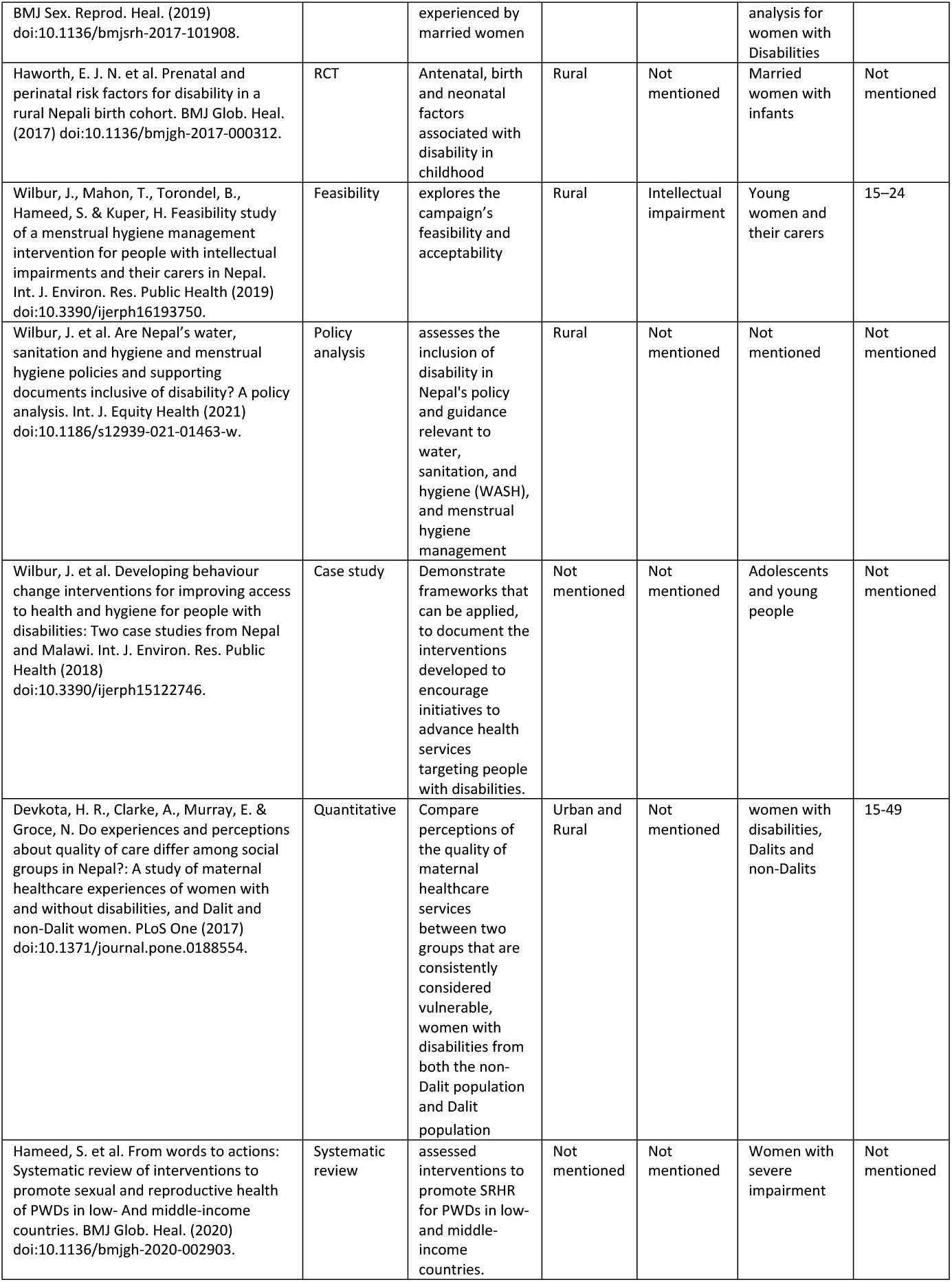
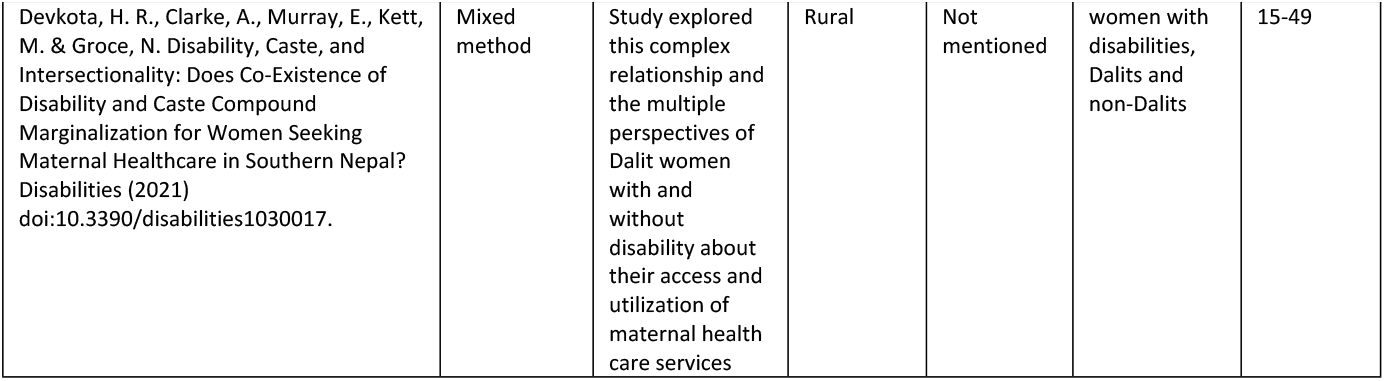
Characteristics of manuscripts included in final review

### Barriers and Facilitators for SRH of PWDs

Out of the 21 publications, eleven reported on individual level(27–35),(36),(37) and institutional and health systems level(27–29,31–34,38,39),(40), seven reported on social and community level(27,29,31,39),(30),(32),(41) and only three reported on structural level factors(31),(42), (38). The overview of the barriers and facilitators experienced by the PWDs are presented in table 2 and 3.

**Table 2:** Overview of the Barriers to SRH for PWDs

| Category | Subcategory | Themes | Number of studies | quotation |
| --- | --- | --- | --- | --- |
| **Individual** | Self-efficacy | Low self-esteem | Four<sup>27,29,31,34</sup> | Many disabled women were embarrassed and this often prevented them from telling their in-laws about their pregnancy<sup>38</sup>. Participants felt humiliated when asking another person to change their menstrual product, and guilt seeing their care providers handle their menstrual blood<sup>29</sup>. |
|  | Education | Knowledge and information | Three<sup>28,29,32</sup> | Nearly 26.7% of PWDs do not have knowledge about the disability rights provided by the government and 94.2% of the respondents think there is no easy access of SRH services for them<sup>28</sup>. |
|  | Socioeconomic status |  | Three<sup>31,32,34</sup> | Those women with disabilities who were ever married are 122 times more likely to utilize SRH services compared to those who were never married<sup>32</sup>. Data suggests that poverty was more of a barrier than embarrassment or bad behavior of health workers, and many women felt it was inevitable that they would deliver at home<sup>38</sup>. |
|  | Severity |  | Five<sup>29,31–33,35</sup> | Women with hearing impairments were identified as being particularly at risk of sexual violence<sup>34</sup>. User's disability status was the influencing factor in receiving services<sup>31</sup>. |
| **Social and community** | Socio cultural norms | Belief and attitude | Four<sup>27,29,31,39</sup> | If a mother cannot see the baby, hear the baby cry or play with them, then what would be the point of having a baby<sup>27</sup>. Disability was also viewed as a curse. Therefore, people feared that they will be doubly cursed if they did not follow restrictions<sup>29</sup>. |
|  | Education | Knowledge and information | Two<sup>27,30</sup>, | Some participants reported that they faced challenges due to preconceived mind-sets and limited understanding about disability and disabled people's desire and expectations<sup>30</sup>. Some of the women with disabilities reported that their parents did not understand their emotional and sexual needs and never talked to them about marriage<sup>27</sup>. |
|  | Support system |  | Three<sup>27,29,33</sup> | In the case where family members are absent, they believe they are more prone to sexual violence, so they provide Depo-Provera injection to prevent the individual from being pregnant<sup>32</sup>. Her husband was blamed for marrying her and excluded for several years by his parents and relatives<sup>27</sup>. |
| **Institutional and health system** | Cost |  | Three<sup>30,33,43</sup>, | Respondents reported to receive good service from government officials only if they have money otherwise there was no service without finances<sup>35</sup>. It is often costly and time-consuming to transport patients to health facilities generally, and it is even more costly for people with disabilities when an additional person is needed to accompany them<sup>31</sup>. |
|  | Social norms | Behavior | Three<sup>29,31,34</sup> | One care provider reported that a teacher sent her daughter home from school at the onset of menarche, and the care provider never sent her back<sup>29</sup>. |
|  |  | Providers gender |  | All the women interviewed reported that they felt uncomfortable while examined by a male provider for maternity care and hesitated to seek services next time<sup>31</sup>. |
|  |  | Attitude |  | Many women with disabilities reported that the attitudes of providers and their understanding about disability were negative and often discouraging, expressing concern about SRH choices of women with disabilities<sup>30</sup>. |
|  | Support from authorities |  |  | Respondents reported that even the authorities'/police officials, when are reported to domestic violence do not provide support to the disabled<sup>35</sup>. |
|  | Resources | Physical (infra structure/ wait time) | Six<sup>28-31,38,39</sup> | Inconvenient and limited opening hours of health facilities were often reported by participants in their qualitative interviews as one of the reasons for non-utilization of services<sup>31</sup>. |
|  | Education | Knowledge | Three<sup>30,33,34</sup>, | The rejection of women with disabilities by the health facility for services could be an indicator of lack of knowledge and experience among providers or established prejudice<sup>31</sup>. |
|  |  | Communication skills |  | Providers did not explain things to them properly or give information about their pregnancy while attending antenatal clinics. It was evident that only a very few were advised to go for delivery in a health facility and none of them were informed about the need for post-natal check-ups<sup>30</sup>. Even in hospitals, they do not get the same treatment as others if they do not have a sign language interpreter. Sometimes, they communicate by writing but it is impossible for illiterate people do that<sup>33</sup>. |
| **Structural** | Policy | Inclusion | One<sup>42</sup> | The act is silent about reproductive and sexual health rights of women/girls with disabilities and management of reproductive impairments explicitly<sup>42</sup>. |
|  | Resources | Implementation and funding | One<sup>31</sup> | Due not only to limited resources but to poor implementation of policies, the most marginalized and vulnerable women are the least likely to receive these benefits<sup>31</sup>. A respondent reported she was asked to pay 60 thousand to access maternal healthcare services in multiple health facility, even though safe motherhood program is in place<sup>31</sup>. |
|  | Organization Capacity |  | One<sup>38</sup> | Despite several organizations working on disability issues in the district, other organization are providing SRH services<sup>38</sup>. |

**Table 3:** Overview of facilitator for SRH and PWDs

| Category | Subcategory | Themes | Number of studies | quotation |
| --- | --- | --- | --- | --- |
| **Individual** | Empowerment | Coping | Three<sup>27,35,38</sup> | Most women had chosen their partner, a stark contrast to the social practices, where arranged marriages remain the norm. When families had not considered arranging a marriage for them; they had sought a partner of their own and lived separately from the extended family<sup>27</sup>. |
| **Social and community** | Social norm | Beliefs | One<sup>27</sup> | Many of the participants believed that emotions and desires about sexuality and pregnancy for women with disabilities are the same as for women without disabilities<sup>27</sup>. |
|  | Social support mechanism | Emotional and financial | Three<sup>27,32,34</sup> | Home-based refugees reported family members, especially mothers, as safe resources. While self-help activities were on the whole limited, participants noted their ability and willingness to share information with each other and with other refugees and leaders in the community<sup>34</sup>. They reported that their partners had married them expecting to acquire their parent's property as part of the dowry, which was an incentive for the marriage<sup>27</sup>. |
| **Institutional and health system** | Social Norm | Attitude | One<sup>30</sup> | Slightly higher mean attitude towards disabled persons scores to those who gave maternal healthcare services to women with disabilities than those who did not (79.49 vs 76.35), however, this difference was not significant $P > 0.05$ <sup>30</sup>. |
|  |  | Respectful |  | Many reported that the providers were kind, caring and treated them respectfully in a welcoming environment. Several stated that they received counselling and advice in their ANC visits and support during childbirth by the nurses and doctors<sup>30</sup>. |
| **Structural** | Organizations for PWDs |  | Two<sup>38,36</sup> | Sample reported presence of several organizations working on disability issues in the district<sup>38</sup>. Another study reported campaign ensuring to help people with intellectual impairment and their carers in regards to menstrual hygiene management<sup>36</sup>. |

#### Individual

Out of the eleven publication reporting on individual level barriers (27–35),(36,40), four publications(28,29,32),(36) identified lack of information and knowledge about the disability and SRH service availability as a barrier. For example, a study exploring barriers to menstrual hygiene among adolescent girls with disabilities reported that the guardian were in disbelief when they had menses as they presumed menarche would be delayed or not have menses(29). A quantitative study exploring challenges of seeking SRH services by PWDs reported that nearly 94.2% thought the SRH services were inaccessible(28), and another article reported that women with disabilities do not report violence because they lack information on where and how to report(33). Four articles identified perceived low self-efficacy and esteem from experiencing constant fear and humiliation as a barrier for women with disabilities to exercise SRH rights (27,29,31,34). Studies reported they were not able to share their needs or problems (31), or even too embarrassed to share the news of their pregnancy to their parents and in-laws (34).

Three publications highlighted socio economic status as a barrier (31,32,34). One study exploring factors affecting utilization of SRH services reported, compared to unmarried women with disabilities those married were more likely to utilize SRH services(31). Similarly, an article reported poverty was a bigger barrier than health worker’s behavior to access maternal health care therefore women with disability often choose to give birth at home(34). Five publications highlighted that the type and severity of disability increased their vulnerabilities (29,31–33,35), for example a study among refugee women reported that those with hearing impairments were more likely to experience sexual violence(32). Another study exploring risk factors of violence among women with disability reported being disabled was the major reason for experiencing violence including rape and the situation gets worse as condition deteriorates(33).

This review also found some facilitators, three of the studies reported individual level facilitators, such as coping and receiving preferential services(27,35,38). One study reported how women with disability were challenging the social norms of arranged marriage by taking matters in their own hand to choose a life partner and living separately from their extended family(27). Another study reported that compared to women with no disability those with disability were more likely to report violence (sexual and physical)(35).

#### Social and community

Five publications reported socio cultural norms as a barrier to SRH rights, particularly for women with disabilities(27,29,31,39),(41). Discrimination and harassment based on erroneous beliefs about PWDs’ sexuality, ability to reproduce and child care were common(27,29,31,39). For example, a qualitative study exploring attitudes and behaviors towards women with disability in rural Nepal reported that family and neighbors constantly questioned disabled women’s desire and ability to raise a child, while their ability to perform other physically demanding household chores was never questioned(27). The same study also reported that societal assumptions that women with disability are powerless weakens and instills fear and insecurity in them. Taboos relating to menstruation and pregnancy outside marriages are common, however two publications reported their repercussions were severe for women with disability(29,32). One of the study exploring utilization of SRH services reported women with disability particularly those with no family members were offered/encouraged to use contraception without proper consent with assumptions they were more likely to encounter sexual violence leading to unwanted pregnancy(31). A study from refugee camp setting reported discrimination against women who became pregnant outside marriage forced some to flee their homes, and others to kill themselves(32).

Two publication reported poor knowledge on disability and SRH needs of PWDs as a barrier(27,30), including among health workers (30). A case study of women with disability reported that they were discouraged to have children fearing the off springs will also be disabled increasing the burden of disability in the society (39).

Three studies reported lack of social support as a barrier for SRHR (27,29,33), for example one study reported that she and her husband were banished by his family for marrying (27), while in another the girl was abused and called names for marrying(33). A study among care takers of PWDs reported they felt isolated and overwhelmed from having to take care of the PWDs on their own (29).

In terms of facilitators, the same study (27) that reported strong social norms as barriers also reported belief that emotion and desires about sexuality were similar for women with and without disabilities were also prevalent and thus they should be encouraged to have children for security and support in old age. Three studies reported support from family and friends was valuable resource’s for PWDs(27,32)^,34^. For example, PWDs felt reassured when they were accepted by and received support from the in-laws(27). However, in some instances parents were over protective hindering PWDs prospect of marriage, while some parents found suitors willing to marry for dowry but such marriages were reported not to end well(27).

#### Institutional and health system

Eleven publications(27–29,31–34,38,39),(37),(40) reported on institutional level barriers. Six of them reported inadequate physical infrastructure (facilities, opening time, remoteness)(28–31,38,39). For example, one study reported lack of ramps for wheelchair access and inconvenient opening hours of the health facilities(30). Five publications identified cost as a barrier(30,33,43),(37),(40)For example, a study reported that good health care services are not accessible for those without resources to pay them(33). Another study among disabled woman in rural setting further elaborated it was not just the users fees, but also travel cost of her and the companion, as they are unable to travel on their own(30).

Inadequate education (knowledge and skills) of health workers to effectively interact and communicate with PWDs was reported by three publications(30,33,34). For example, a study focusing on healthcare provider’s attitudes(38) highlighted health care providers avoid talking to the PWDs when providing care and lacked confidence to handle maternal health care such as child birth. Health facilities often denied health care to persons with severe disabilities citing incapability to provide services(30).

Three publications(29,31,34) highlighted social norms such as belief and attitudes help by service providers driving health system barriers. For example, a study reported that doctors advised women with disability to not to have any more children with no further explanation(38). According to one qualitative study among adolescent girls with disability(29), adolescent girls were sent home by teachers after having menses never returning to school, such attitude and response by teachers ended any chance they had of receiving SRH education from school. Similarly, two study(29,33) reported lack of support by those in authority, a study focusing on violence against woman with disability showed that police officials do not provide any kind of support when violence is reported(33). The same study further elaborated that violence were less likely to be reported if it was inflicted by an intimate partner(33).

Only one publication reported institutional facilitator that the health workers providing maternal health services to women with disability had better attitude compared to those not providing services to women with disability, in support the women with disability also shared they felt respected by health care providers(30).

#### Structural

The findings for structural barriers are based on findings from article review as well as the review of policies and data sources. Supplementary table 2 shows the list of policy documents (strategies, guidelines, plans and acts) the researcher reviewed. Altogether 19 policies (10 on broader health policies, 9 specific to SRH and 2 on protecting rights of the PWDs) of Nepal government were reviewed.

With regards to structural barriers, this review found little inclusion of SRH rights of PWDs in policies and programming. While the constitution of Nepal guarantees universal, non-discriminatory and free SRH care services for marginalized groups including PWDs(42), these aspirations were not carried through into policy. The National Health Policy 1991 mentions engaging with the private sector to provide rehabilitation services for PWDs whereas the most recent National Health Policy 2019 talks of providing reproductive health services through lifecycle approach in providing with no particular mention of PWDs. Another policy analysis document shows that disability is not fully covered in policy documents and MHM’s disability policy commitments are almost non-existent(41), this resulted in limited professional understanding of the issues, as service providers had no training.

Only eight out of 19 policy documents reviewed touched upon SRH rights of PWDs, but most mentioned PWDs as a marginalized group without outlining specific activities or strategies designed to meet their needs. Fig 2 shows that the timelines of policies that have included SRH rights of PWDs, almost all of them are after the CRDP. Two acts(44,45) and a guideline(46) have called for protecting SRH rights of PWDs with special focus on women through provision of disability friendly reproductive health services including sexual harassment and violence. The guideline(46) further elaborated the need to raise awareness and enhance the communication and counselling skills of health workers and to raise awareness among PWDs on availability of SRH services. A standard for maternal and newborn care(47) recommends equitable access to perinatal care for women with disabilities stressing the need for additional information and resources to facilitate the labor, child birth and lactation. Similarly, Nepal safer motherhood and newborn health roadmap talks about improving access to maternal and newborn care through home visits to provide care and provision of devices to facilitate communication. A costed implementation plan(48) for family planning allocated funds to develop a national strategy to address legal and socio-cultural barriers faced by PWDs in accessing family planning services.

**Fig 2.**
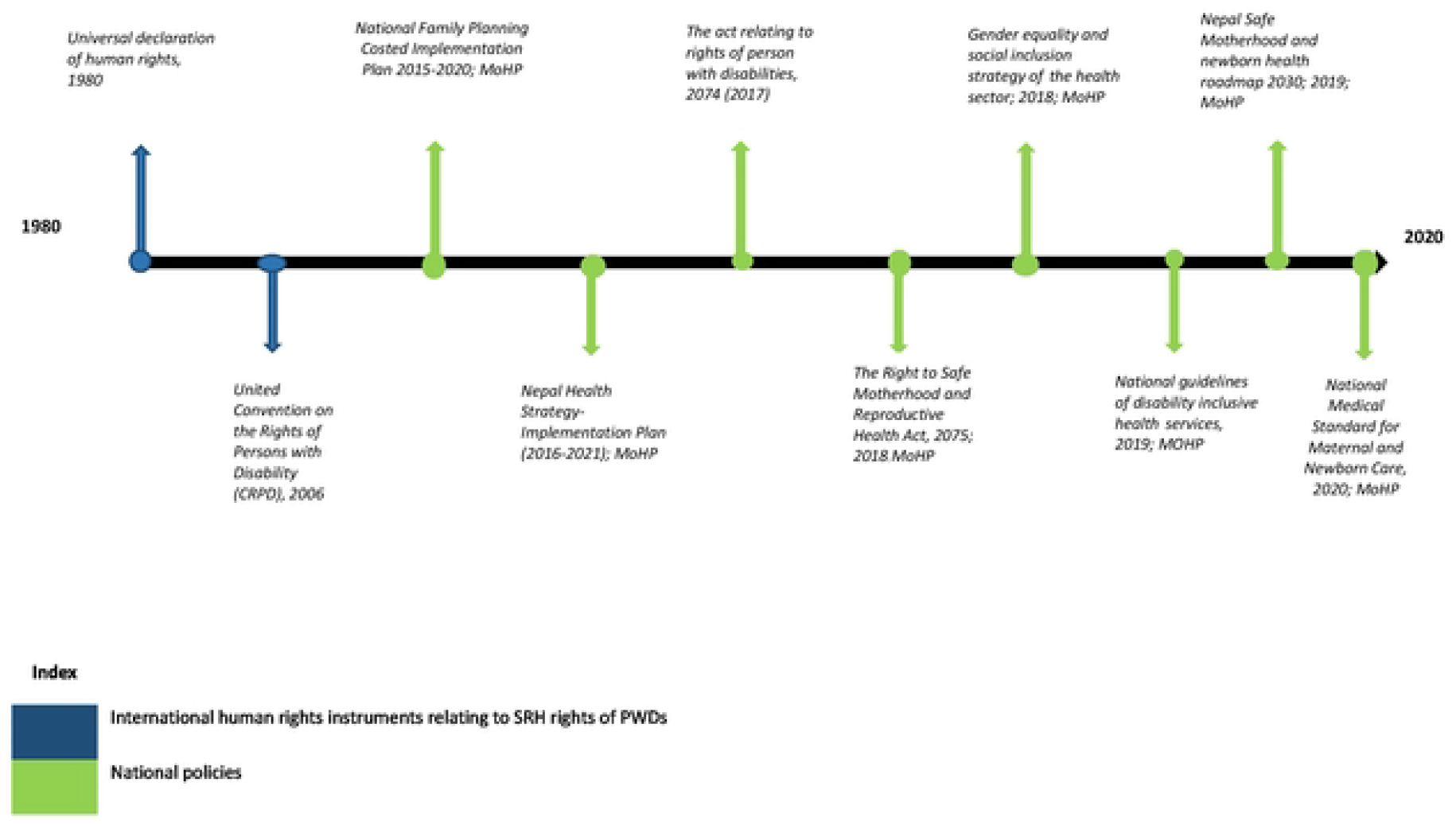
Timeline of policy documents that have mentioned SRH of PWDs

Two of the policies(46,49) reviewed urged the MoHP to conduct research assessing the SRH needs of PWDs and one(49) of them further calls to establish a surveillance system for PWDs in reproductive age and share the information with stakeholders to guide evidence based policy and programming for PWDs. However, we found paucity of data on SRH of PWDs to guide development of SRH programs for PWDs. Supplementary Table 3 show national level data sources(50–57) (1 census, 6 surveys and 1 health system monitoring data). Four of them collected data on SRH, such as utilization of SRH services(50–52,55), knowledge and access to SRH services(50,51,55) and violence and harassment experience (50,51). However, only two data sources census and multi-indicator cluster survey had data disaggregated by disability status, but only the later had SRH data on PWDs disaggregated by gender.

Our findings from assessment of websites of 12 national level organizations that the authors knew worked for PWDs are presented in supplementary table 4. We found SRH rights and services were not prioritized within their programming, only two organizations mentioned taking up SRH issues of PWDs. However, it was not clear what kind of interventions were implemented and our searches yielded no published work about them or on any other interventions hinting lack of SRH intervention for PWDs. This was validated from one of the publication(34) included in the review that reported organizations working with PWDs did not provide SRH interventions (information and services). Similarly, a study(29) exploring barriers to menstrual hygiene among women with disabilities reported unavailability of support mechanisms or networks for PWDs and their care takers.

## Discussion

This paper assessed evidence on barriers and facilitators faced by PWDs in recognizing their rights and accessing SRH services in Nepal. The common barriers reported in the reviewed literature were exclusion of PWDs from health and SRH policies, lack of knowledge, socio cultural norms and inadequate support system. However, facilitators to PWD uptake of SHR services were rarely discussed – they were and only reported in passing by five studies. Overall, many of the barriers were also reported in studies from low and middle income countries in Sub-Saharan Africa(58), rural Cambodia(59), and North India (60) and the UK(61).

The literature on the situation in Nepal suggests similarities with the situation in other countries: lack of knowledge and education (58–63), lack of disability friendly infrastructure(58) and poor attitude of health care providers (58,60,62). Barriers to SRH for PWD in Nepal that we identify in our review are similar to barriers to access among PWD for general health care(64). We found inadequate mention of PWDs and their rights in health policies in Nepal, in fact health policies appear to have regressed in terms of highlighting rights to healthcare for PWDs. For instance, while the 1991 national health policy mentions providing rehabilitative services for PWDs, the 2019 policy omits this altogether. Such systematic exclusion of PWDs in the sectoral plans have also been reported in reviews from Africa and Mediterranean regions(58,65,66). If left unchallenged the SRH rights of PWDs will continue to be neglected undermining the country’s ability to ensure universal access to SRH services. It is worth noting that some of the most recent policy documents have begun to fulfill the country’s commitment to international human rights instruments by including SRH needs of PWDs. Despite these policy initiatives, a study among women with disability reported that poor implementation of policies, and cost of the maternal health care were major hindrance to access SRH care(30). Thus, raising questions on the level of commitment, and highlighting the need for further exploration of gaps in policy implementation.

In parallel with our findings a systematic review including studies mostly from high income countries reported negative attitudes and stigma around SRH needs of women with disabilities, it found people including parents were uncomfortable with PWDs being sexually active(67). In fact, the PWDs themselves held such misconceptions often due to inadequate knowledge about their SRH rights, poor awareness on policies and provisions made by the government (68) leading to reluctance in seeking and demanding SRH information and services. Similarly, we found girls with disabilities were discouraged to go to school after onset of menses, as was also reported by a systematic review outside Nepal depriving girls from school was a coping mechanisms parents used to avoid embarrassment caused by their daughter’s inability to follow social and cultural norms on mensuration(67). These misconceptions imply poor understanding and awareness of disability and sexuality of PWDs; therefore, this suggests that educating the public on SRH needs of PWDs and their right to lead a safe and satisfying sexual life. Such education can also be mainstreamed into the formal education system for lasting positive socio-cultural change. In addition, activities to empower persons living with disabilities such as educating them on their SRH rights are important for them- to confidently access SRH services.

Sexual violence and subsequent lack of access to legal and social support for women with disabilities found during this review have also been reported by a study from Senegal(69). A probable reason for deterring them from seeking care and support may be due to fear of victimization.(17) To encourage women with disabilities to report violence and use SRH services, the care and support system may have to shift from imposing long term contraception on women with disabilities to avoid the consequences of sexual violence by investing in training and education of the social and health care workers to recognize signs of violence and respecting women with disabilities and encouraging them to understand their rights. In addition, those providing care or guardians must also be empowered to dismiss wrong socio-cultural beliefs, one way to achieve this is to form support groups of guardians, where they can discuss their concerns, find solutions and voice their concerns jointly when dealing with authority.

Several health system barriers such as lack of knowledge about needs of PWDs, poor communication skills of health care professionals, and cost of health care identified during this review undermine the SRH rights of PWDs. Several studies (62,70,71) have shown these factors negatively affect the relationship between health care providers and PWDs discouraging them from using health care services. Therefore, for effective interpersonal relationship based on clear communication and trust, the government should provide regular training to the health care providers on SRH rights of PWDs and to provide non-judgmental SRH information and services. Similarly, health facilities must accommodate the specific needs of persons with all forms of disabilities such as making provisions for an in house or outsource when needed a sign language interpreter, wheelchair accessible ramps, toilets, beds, and equipment.

Agreeing with our findings, a systematic review from low and middle income countries has reported a strong link between poverty and disability (72), while others have reported type and severity of disability (62,70) determined access to health care. Poverty makes SRH services unaffordable, and also the negative attitudes towards PWDs may get worse with poverty(72). Such findings underscore the urgent need to elaborate on the right based equitable health service vision inscribed in the policy documents with sufficient allocation of resources to implement them.

Apart from barriers, our study also identified few facilitators such as PWDs were empowered enough to choose their own partner, supportive family members, and instances of good treatment by health care providers. These show that social support system exists in the society, there is the need to identify and tap on such good examples to develop a well thought out culturally appropriate programs and interventions to support PWDs and their family to exercise their SRH rights in respectful manner.

Overall, our review has two important strengths. First, to the best of our knowledge, this is the first review to identify and synthesize the current literature on perceived barriers and facilitators to SRH among PWDs and the interventions to address their needs in Nepal. Second, we reviewed policies, published paper and national sources of data to enrich our findings from various angles. However, this review also had some limitation, first, restricting our search to only one database and studies published between 2011-2021 may have excluded some relevant publications. Second, this review did not report on the extent of interaction between these barriers to influence access to SRH services. Third, most of the studies reviewed were done among women with disabilities, therefore the barriers and facilitators faced by men are not properly articulated in this review.

## Conclusion

In summary, we found that inadequate inclusion of PWDs in health sector policies, lack of knowledge about SRH needs, misconception and poor attitude and lack of social support system were the main barriers to accessing SRH rights and services in Nepal. Understandings from this review could be used to develop a roadmap to address access barriers at various level; national, institutional or health system, individual and community level. In addition, further studies exploring experience from different subgroups of PWDs, their guardians and health care providers are needed to substantiate the findings from this review.

## Data Availability

As this was a scoping review - Data analyzed in the current research were review of the existing literatures in google search engine and pubmed database

## Acknowledgement

The authors would like to thank Catherine McGowan for guiding with the scoping review process.

## Funding

The authors declare that no funds, grants, or other support were received during the preparation of this manuscript.

## Declaration

We confirm that the manuscript has been read and approved by all named authors and that there are no other persons who satisfied the criteria for authorship but are not listed.

## Conflict of Interest

All authors declare that they have no conflicts of interest.

## Notes

### Competing Interest Statement

The authors have declared no competing interest.

### Author Declarations

As this was a scoping review - ethical approval was not needed for the research conducted

